# DPP9 deficiency: an Inflammasomopathy which can be rescued by lowering NLRP1/IL-1 signaling

**DOI:** 10.1101/2021.01.31.21250067

**Authors:** Cassandra R. Harapas, Kim S. Robinson, Kenneth Lay, Jasmine Wong, Ricardo Moreno Traspas, Nasrin Nabavizadeh, Annick Raas-Rothschild, Bertrand Boisson, Scott B. Drutman, Pawat Laohamonthonkul, Devon Bonner, Mark Gorrell, Sophia Davidson, Chien-Hsiung Yu, Hulya Kayserili, Nevin Hatipoglu, Jean-Laurent Casanova, Jonathan A. Bernstein, Franklin L. Zhong, Seth L. Masters, Bruno Reversade

**Author notes:** Joint senior authors to whom correspondence should be addressed: Franklin Zhong, Seth Masters, Bruno Reversade. These authors contributed equally to this work (Cassandra R. Harapas, Kim S. Robinson, Kenneth Lay).

## Abstract

Dipeptidyl peptidase 9 (DPP9) is a direct inhibitor of NLRP1, but how it impacts inflammasome regulation in vivo is not yet established. Here, we report two families with immune-associated defects, skin pigmentation abnormalities and neurological deficits that segregate with biallelic *DPP9* rare variants. Using patient-derived primary cells and biochemical assays, these variants are shown to behave as hypomorphic or loss-of-function alleles that fail to repress NLRP1. Remarkably, the removal in mice, of a single copy of either *Nlrp1a/b/c, Asc, Gsdmd, Il-1r*, but not *Il-18*, was sufficient to rescue the lethality of *Dpp9* mutant neonates. These experiments suggest that the deleterious consequences of DPP9 deficiency are mostly driven by the aberrant activation of the canonical NLRP1 inflammasome and IL-1β signaling. Collectively, our results delineate a Mendelian disorder of DPP9 deficiency driven by increased NLRP1 activity as demonstrated in patient cells and in a mouse model of the disease.

## Main

The human NLRP1 inflammasome is a multi-protein complex that is triggered by viral infections^1,2^ or danger-associated signals^3^, resulting in the release of proinflammatory cytokines, including IL-1 family members, IL-1β and IL-18^4^. Dysregulation of the human NLRP1 inflammasome is associated with a range of inflammatory Mendelian diseases such as Multiple Self-healing Palmoplantar Carcinoma (MSPC, MIM615225)^5^, Juvenile Recurrent Respiratory Papillomatosis (JRRP, 618803MIM)^6^, and Autoinflammation with Arthritis and Dyskeratosis (AIADK, MIM617388)^7^. In the absence of any cognate triggers, inflammasome sensors in the family of Nod-like receptors (NLRs) are maintained in an inactive state by a number of regulatory proteins. For example, NLRP1 is bound to, and repressed by, the enzymatic activity and scaffolding function of dipeptidyl peptidase 9 (DPP9) and DPP8^7,8^. The role of DPP9 as a negative regulator of human or mouse NLRP1 has been established through pharmacological inhibition of DPP9 and inactivation of DPP9 in cell lines in vitro, but its function has not been documented in vivo.

As part of a cohort of patients with inborn errors of immunity, we studied a child (F2: II.4) with signs of susceptibility to infections consisting of recurrent facial HSV lesions and airway inflammation, including chronic intermittent asthma and primary pulmonary tuberculosis. Exome sequencing disclosed a germline homozygous stop-gained mutation (c.2551C>T; p.Q851*) in *DPP9* (<u>MIM608258</u>) (Fig. 1a, b, e) which was heterozygous in the parents. A second proband, a 6-year-old child (F1: II.2), was identified with help of the GeneMatcher^9^ database and carried compound heterozygous *DPP9* variants with a paternal missense mutation (c.449G>A, p.G167S) and a maternal early truncating mutation (c.641C>G; p.S214*) (Fig. 1a-c). Both children shared a triad of symptoms summarized in Table 1 which partially overlapped with NLRP1 gain-of-function mutations causing AIADK^7^. This consisted of a general failure to thrive, skin manifestations and blood anomalies, which in the case of F1: II.2 required hematopoietic stem cell transplant. The stop-gain mutations found in the two probands are expected to completely abolish the enzymatic activity of DPP9, as its catalytic domain is situated at the C-terminus. The missense mutation p.G167S found in proband F1: II.2 affects a residue in a flexible loop that is located at the periphery of the DPP9 substrate binding site (Fig. 1d). Thus, the DPP9^G167S^ mutation is predicted to diminish the substrate binding and/or catalytic efficiency. To date, no homozygous damaging variants have been reported for *DPP9* which according to genomAD is significantly averse to loss-of-function mutations^10^.

**Fig. 1:**
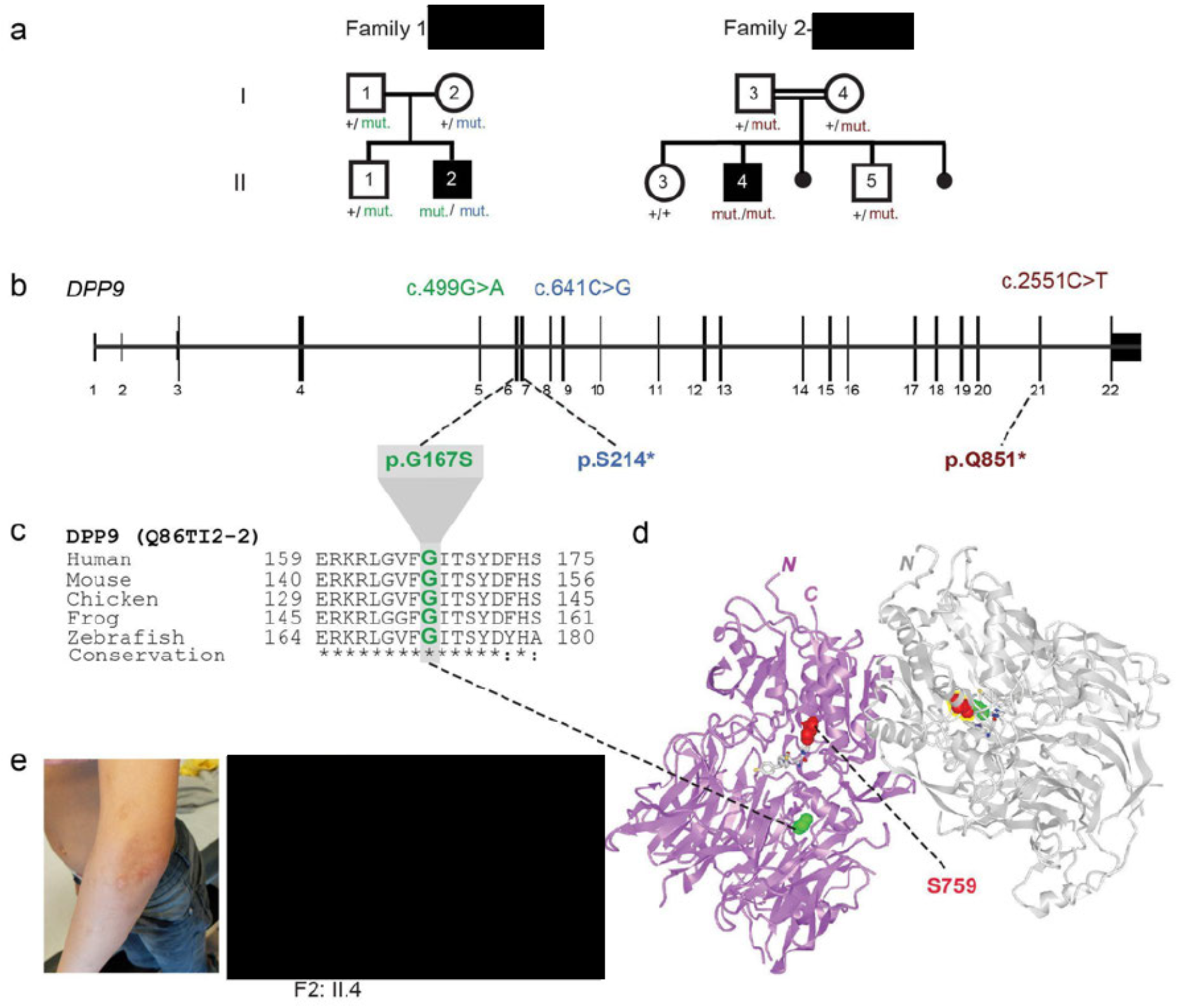
Two families segregating recessive loss-of-function *DPP9* germline variants. **a**, Pedigrees of two families harbouring *DPP9* mutations. Proband from Family 1 is compound heterozygous, with paternal p.G167S and maternal p.S214* germline mutations. Proband from Family 2 carries a homozygous recessive p.Q851 * mutation. **b**, Exon-intron genomic organization of *DPP9* with positions of identified germline mutations. **c**, Phylogenetic conservation of the Gly167 residue, which is mutated in Family 1, across vertebrate species. **d**, Protein structure of a DPP9 homodimer (PDB ID: 6EOR) depicting location of p.G167 on the periphery of the substrate binding site and p.S759 in the catalytic domain. **e**, Photographs of proband from Family 2 (II.4) depicting recurrent HSV facial lesions, milia, and skin pigmentation anomalies on his arms.

**Table 1.** Syndrome caused by biallelic Loss-of-Function *DPP9* mutations.

|  |  |  |  |  |
| --- | --- | --- | --- | --- |
| *Gene* | HPO<br>Terms | *DPP9* (MIM608258) |  |  |
| **Clinical synopsis and genetics** |  |  |  | **Present in both probands** |
| Country of origin |  |  |  |  |
| Propositus number<br>(refer to pedigrees) |  | **II:1** | **II:3** |  |
| Gender, (year of birth) |  | Male | Male |  |
| IUGR, (birth weight in kg) | HP:0001511 | **+ (2.3)** | **- (3.6)** |  |
| Autosomal recessive inheritance | HP:0000007 | compound heterozygous | homozygous | ✓ |
| cDNA change<br>(MutationTaster prediction) |  | maternal:<br>c.641C>G<br>paternal:<br>c.499G>A | c.2551C>T |  |
| Expected protein change (Q86TI2-2) |  | maternal:<br>p.S214*<br>paternal:<br>p.G167S | p.Q851* |  |
| Observed protein change (Q86TI2-2) |  | maternal: p.0<br>paternal:<br>p.G167S | p.0 | ✓<br>Loss-of-function mutations |
| **Craniofacial dysmorphisms** |  |  |  |  |
| Downslanted palpebral fissures | HP:0000494 | + | + | ✓ |
| Broad forehead | HP:00002373 | + | - |  |
| **Bone / skeletal abnormalities** |  |  |  |  |
| Proportionate short stature | HP:0003508 | + | + | ✓ |
| **Immune defects** |  |  |  |  |
| Infantile eczema | HP:0000964 | + (after BM transplantation) | + | ✓ |
| Recurrent fevers | HP:0032323 | - | *bimonthly* |  |
| Asthma | HP:0002099 | - | + |  |
| Repeated infections | HP:0002719 | + | + | ✓ |
| Herpes infections | HP:0005353 | - | + (*facial*) |  |
| Bronchitis | HP:0012<br>387 | + | + | ✓ |
| Prone to acute otitis media | HP:0000<br>403 | + | + | ✓ |
| Pancytopenia | HP:0001<br>876 | + | + | ✓ |
| Anemia | HP:0001<br>903 | + | + | ✓ |
| Selective immunodeficiency | HP:0002<br>721 | + | + | ✓ |
| Severe allergies | HP:0012<br>393 | + (*sesame*) | + | ✓ |
| **Behavioural/Neurological traits** |  |  |  |  |
| Learning Disability | HP:0001<br>328 | + | + | ✓ |
| Speech delay | HP:0000<br>750 | + | + | ✓ |
| Slurred speech | HP:0001<br>350 | + | + | ✓ |
| Autism spectrum disorder | HP:0000<br>729 | + | - |  |
| **Skin findings and its appendages** |  |  |  |  |
| Thick skin | HP:0001<br>072 | + | - |  |
| Anhidrosis | HP:0000<br>970 | - | + |  |
| Dry skin | HP:0000<br>958 | - | + | ✓ |
| Atopic dermatitis | HP:0001<br>047 | + | + | ✓ |
| Petechiae | HP:0000<br>967 | + | + | ✓ |
| Fair hair | HP:0002<br>286 | + | + | ✓ |
| Premature hair<br>graying | HP:0002<br>216 | - | + |  |
| Hypo- and<br>hyper-pigmented<br>macules | HP:0007<br>441 | + | + | ✓ |
| Delayed wound<br>healing | HP:0001<br>058 | - | + |  |
| **Other clinical**<br>**manifestations** |  |  |  |  |
| Failure to thrive | HP:0001<br>508 | + | + | ✓ |
| Febrile seizures | HP:0002373 | + | - |  |
| Cryptorchidism /<br>hypospadias | HP:0000028 | + | - |  |
| Inguinal hernia | HP:0000776 | + | - |  |
| Poor feeding | HP:0011968 | + | + | ✓ |

**n.d., not determined; +, affirmative; -, negative; BM, bone marrow**

To investigate the impact of the *DPP9* variants on mRNA expression and enzymatic function, primary skin cells were isolated from affected patients of both families (Fig. 2a). When compared with control primary fibroblasts, and those derived from the heterozygous parents of family 2 (F2: I.3 and I.4), a significant decrease in DPP9 protein was found in the proband’s (F1: II.2) fibroblasts. This was consistent with one allele not encoding the full-length protein due to an early stop mutation p.S214* (Fig. 2a, lanes 1 and 2). Notably, we could not observe any endogenous DPP9 protein in the proband of family 2 (F2: II.4), indicating that the stop-gain mutant (p.Q851*) was degraded via the nonsense-mediated decay pathway (Fig. 2a, lanes 1 and 5). Thus, the p.Q851* *DPP9* variant probably yields a loss-of-expression, resulting in a protein-null allele. We next sought to assess the enzymatic activity of the DPP9^G167S^ variant towards a model substrate Gly-Pro-AMC (Fig. 2b, Extended Data Fig. 1a). The p.G167S mutation was found to reduce the enzymatic activity to ∼15% of that of wild-type DPP9.

**Fig. 2:**
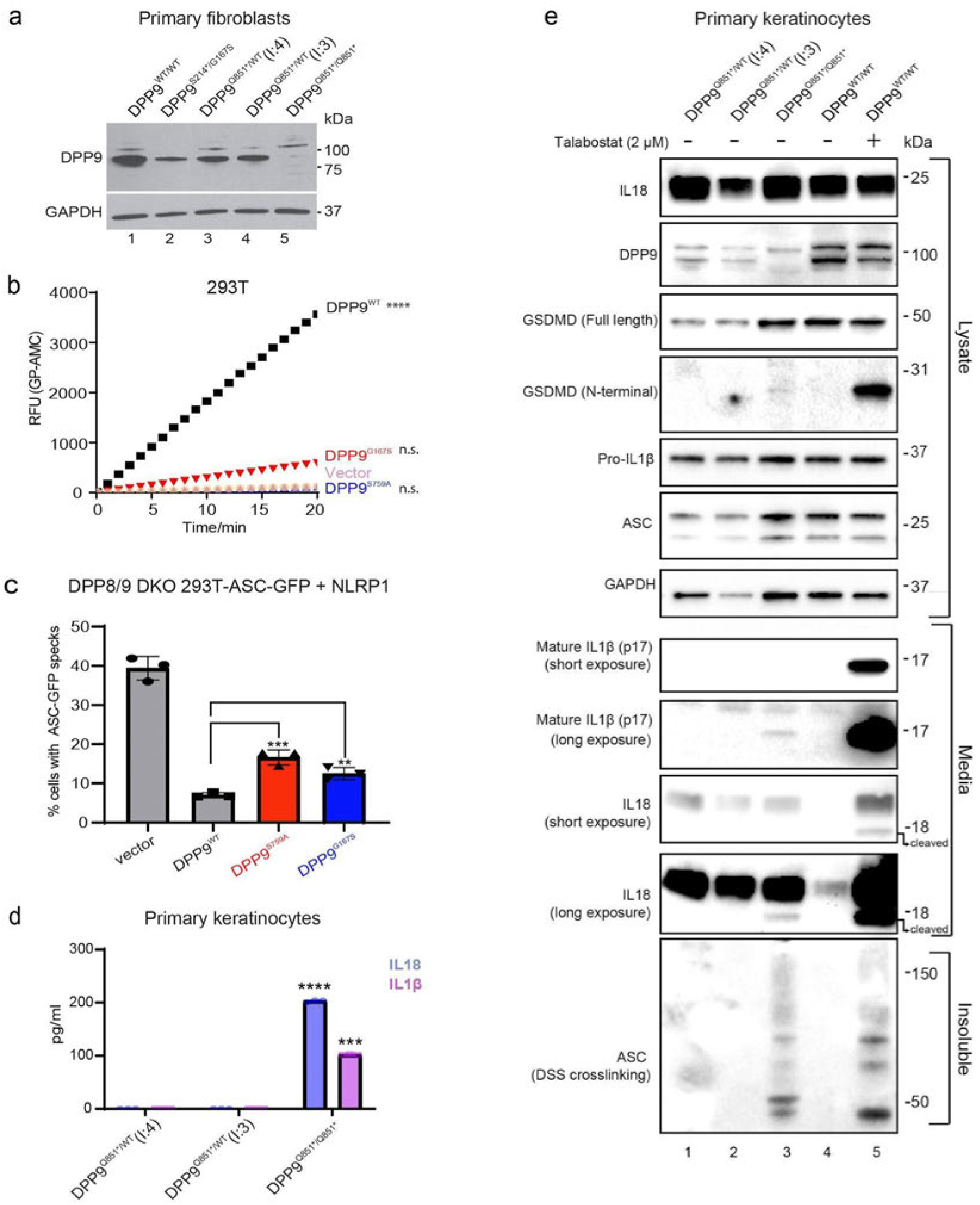
Patients’ cells lack DPP9 or produce a strong enzymatic hypomorph. **a**, Western blot analysis shows reduction of DPP9 protein expression in human dermal fibroblasts isolated from family 1 and family 2 probands. Western blot analysis of DPP9 in primary dermal fibroblasts of an unrelated control (WT/WT) (lane 1), affected individual from family 1 (proband (II:2) S214*/G167S) (lane 2), two unaffected individuals from family 2 ((I:4) and (I:3) Q851*/WT) (lane 3 and 4 respectively) and affected individual from family 2 (proband (II:4) Q851*/Q851*) (lane 5). **b**, DPP8/9 enzyme assay shows a reduction in DPP8/9-derived enzymatic activity in DPP9 S759A and G167S mutants. 293T cells were transfected with either vector, wild-type DPP9, DPP9 S759A or DPP9 G167S and lysed in PBS 1% Tween 20, 48 h after transfection. 0.3 µg of total lysate was then incubated with Gly-Pro-AMC fluorescence substrate. AMC fluorescence was measured for 20 mins at 25 °C in a 50 µl reaction every minute on a spectrometer. One-way ANOVA. ****P<0.001 calculated by comparing the vector control to wild-type DPP9, DPP9^S759A^ or DPP9^G167S^. **c**, DPP9 S759A and G167S mutants have decreased ability to repress NLRP1 inflammasome. 293T-ASC-GFP DPP8/9 DKO cells were transfected with a vector control or WT NLRP1 together with wild-type DPP9, DPP9 S759A, or DPP9 G167S. The percentage of cells with ASC-GFP specks were counted in three different fields after fixation. **P<0.01, ***P<0.001 (one-way ANOVA). n=3 transfections. More than 100 cells were scored per condition. **d**, Family 2 proband primary human keratinocytes experience constitutive inflammasome activation. IL1β and IL18 ELISA of the culture supernatant from primary keratinocytes from the two unaffected individuals from family 2 ((I:4) and (I:3) Q851*/WT) and affected individual from family 2 (proband (II:4) Q851*/Q851*). ****P<0.001, ***P<0.01 (one-way ANOVA). **e**, Family 2 proband (II:4) primary human keratinocytes experience constitutive inflammasome activation. Western blot analysis of inflammasome components in primary keratinocytes of two unaffected individuals from family 2 ((I:4) and (I:3) Q851*/WT) and the affected individual from family 2 (proband (II:4) Q851*/Q851*). Additionally an unrelated control (+/+), treated with or without the potent DPP8/9 inhibitor Talabostat (2 µM) was included as a positive control for NLRP1 inflammasome activation.

Given that DPP9 has been identified as a specific interacting partner and inhibitor of human and mouse NLRP1^7,8^, we measured the ability of DPP9^WT^, catalytically-dead DPP9^S759A^ or the patient DPP9^G167S^ mutant to repress NLRP1 when re-expressed in DPP8/DPP9 double KO 293T-ASC-GFP reporter cells (Fig. 2c, Extended Data Fig. S1b, c). Exogenous DPP9^WT^ repressed NLRP1-dependent ASC-GFP speck formation. In contrast, both DPP9^S759A^ and DPP9^G167S^ were less effective in inhibiting ASC speck formation. Taken together, these experiments suggest that these DPP9 patient variants are less able to maintain NLRP1 in the inactive state.

We and others have demonstrated that NLRP1 is the most prominent inflammasome sensor in human keratinocytes and that these cells readily undergo pyroptotic cell death in response to NLRP1 activation^5,11^. Using primary keratinocytes isolated from family 2 and the Luminex platform, we found that the cytokine and chemokine profile of the proband differed from those of his heterozygous parents (Extended Data Fig. 1d). We demonstrated that the proband keratinocytes, as do keratinocytes from patients with gain-of-function NLRP1 mutations^5^, spontaneously secreted active IL-1β and IL-18 into the media without any exogenous stimulus. Despite measurable IL-1 release visualised by ELISA and western blot (Fig. 2d, e respectively), the patient’s keratinocytes were viable in culture and could be serially cultured (Extended Data Fig. 1e) for at least 4 passages. This cytokine release was also accompanied by ASC oligomerization, as demonstrated using a DSS crosslinking assay, and GSDMD cleavage (Fig. 2e). These data are consistent with a small fraction of the cells undergoing spontaneous inflammasome activation, without overt pyroptosis. A similar phenomenon has been documented in murine macrophages^12^.

To dissect the pathogenicity of the loss of DPP9 in vivo, we took advantage of the DPP9 mutant mouse line which carries the catalytic-dead DPP9 p.S729A mutation. Homozygous *Dpp9*^*S729A/S729A*^ knock-in mice die within 24 hours of birth^13,14^. Based on our results obtained in patient-derived primary cells, we examined whether this lethality might be associated with overactivation of the NLRP1 inflammasome. Indeed, homozygous loss of all *Nlrp1* alleles (*Nlrp1a/b/c*, herein referred to as *Nlrp1*) protected *Dpp9*^*S729A/S729A*^ mice from neonatal lethality. Furthermore, deletion of even a single *Nlrp1* allele was also sufficient to generate viable animals (Fig. 3a, Extended Data Fig. 2a, b). *Dpp9*^*S729A/S729A*^*Nlrp1*^*-/-*^ mice were healthy and proved to be fertile, although runted compared to *Dpp9*^*S729A/+*^*Nlrp1*^*-/-*^ siblings (Fig. 3a, b). Short stature of *Dpp9*^*S729A/S729A*^*Nlrp1*^*-/-*^ was associated with decreased hip width but relatively conserved limb width (Fig. 3c, d).

**Fig. 3:**
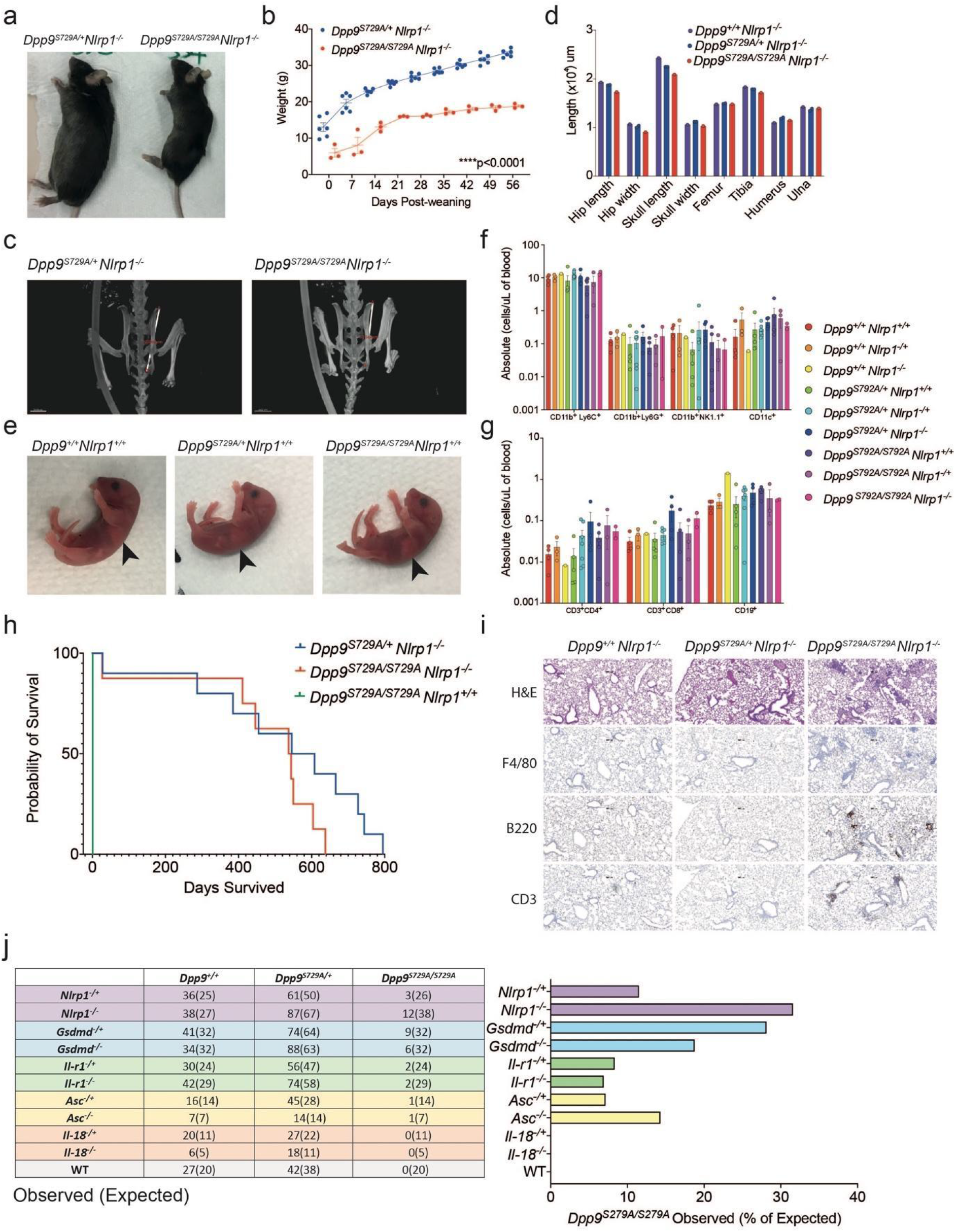
Deficiency of NLRP1 and downstream signaling molecules rescues neonatal lethality of *Dpp9*^*S729A/S729A*^ mice. **a**, A viable, 6-month-old male *Dpp9*^*S729A/S729A*^ *Nlrp1*^*-/-*^ mouse is runted compared to a *Dpp9*^*S729A/+*^ *Nlrp1*^*-/-*^ littermate. **b**, *Dpp9*^*S729A/S729A*^ *Nlrp1*^*-/-*^ mice exhibit stunted weight gain compared to *Dpp9*^*S729A/+*^*Nlrp1*^*-/-*^ controls. Male *Dpp9*^*S729A/S729A*^ *Nlrp1*^*-/-*^ and *Dpp9*^*S729A/+*^ *Nlrp1*^*-/-*^ mice were weighed weekly for 8 weeks post weaning (n=4-5 per genotype). **c**, Decreased stature is associated with decreased hip length but relatively conserved limb measurements, example images of skeletal measurements and **d**, quantification of 6-month-old *Dpp9*^*S729A/S729A*^ *Nlrp1*^*-/-*^ and *Dpp9*^*S729A/+*^ *Nlrp1*^*-/-*^ mice imaged via MicroCT scanning. **e**, Newborn *Dpp9*^*S729A/S729A*^ appear visually normal. Images of 3-6 hour-old pups, black arrows indicate presence of milk spots. **f, g**, FACs analysis of immune cells collected from peripheral blood from 3-6 hour-old pups reveals normal cell numbers from *Dpp9*^*S729A/S729A*^ mice. **h**, Post-weaning, the viable *Dpp9*^*S729A/S729A*^ *Nlrp1*^*-/-*^ mice show no difference in survival compared to *Dpp9*^*S729A/+*^*Nlrp1*^*-/-*^ mice in a Kaplan Meier survival curve, until around 500 days of age (n=8-13 per genotype). **i**, *Dpp9*^*S729A/S729A*^ *Nlrp1*^*-/-*^ display immune cell infiltrate in lungs. Representative section of lung tissue from 6-month-old *Dpp9*^*S729A/S729A*^ *Nlrp1*^*-/-*^ and *Dpp9*^*S729A/+*^ *Nlrp1*^*-/-*^ mice stained with Hematoxylin and eosin or immunohistochemistry of F4/80, B220 or CD3 antibodies. (n=3 per genotype). **j**, Genetic analysis of offspring at weaning age was performed from *Dpp9*^*S729A/S729A*^ crossed to *Nlrp1*^*-/-*^, *Gsdmd*^*-/-*^, *Ilr-1*^*-/-*^ *Asc*^*-/-*^ showing rescue, cross to *Il-18*^*-/-*^ mice did not confer rescue. Error bars represent mean ±SEM. Statistical significance for weight data was determined by a paired t-test. Statistical test for the survival curve was determined by a Mantel-Cox test.

At birth (day 0), *Dpp9*^*S729A/S729A*^ mice appeared normal and were suckling, as evidenced by a milk spot (Fig. 3e). As both DPP9 deficient patients displayed pancytopenia, we used flow cytometry to characterise immune cell populations in the blood of *Dpp9*^*S729A/S729A*^ mutant mice sufficient, heterozygous or deficient for NLRP1, along with relevant controls. Numbers of CD11b+ Ly6C+, CD11b+ Ly6G+, CD11b+ NK1.1+, CD11c+, CD3+ CD4+, CD3+ CD8+ and CD19+ cells in the bone marrow were comparable between all genotypes assessed (Fig. 3f, Extended Data Fig.2c). This is consistent with viable fetal liver chimeras that have previously been generated from this line^15,16^. Similarly, numbers of the same immune cell populations collected from peripheral blood were also comparable between all genotypes accessed (Fig. 3g, Extended Data Fig. 2d). Thus, we do not observe significant haematological abnormalities similar to those observed in patients with germline loss of DPP9 activity.

In order to determine NLRP1-independent effects of DPP9 loss of function we further interrogated the *Dpp9*^*S729A/S729A*^ *Nlrp1*^*-/-*^ line. Aside from being runted, *Dpp9*^*S729A/S729A*^ *Nlrp1*^*-/-*^ mice had a slight decrease in life expectancy (Fig. 3h) compared to control lines. *Dpp9*^*S729A/S729A*^ *Nlrp1*^*-/-*^ mice also displayed immune infiltrate in the lungs which contained foamy macrophages (F4/80+), B cells (B220+) and T cells (CD3+) post 6 months of age (Fig. 3i, Extended Data Fig. 3a, b). While histological analysis of other organs and automated blood cell analysis showed no overt differences between genotypes (Extended Data Fig. 3c-f). Overall, these results suggest inflammasome-independent roles of DPP9 relating to stature and lung inflammation, which is consistent with the broad range of processes regulated by DPP9. Decreased stature and lung inflammation were also recorded in DPP9-deficient patients (Table 1). Notably, two GWAS studies have implicated germline DPP9 variants in the development of pulmonary fibrosis^18,19^ and severe COVID19 pathology^20^.

To further dissect the contribution of downstream components of the NLRP1 signalling axis, we next crossed *Dpp9*^*S729A/S729A*^ mice with strains deficient for effectors of the NLRP1 inflammasome. Both heterozygous and homozygous loss of *Asc, Gsdmd* and *Il-1r* rescued *Dpp9*^*S729A/S729A*^ neonatal lethality to a similar extent as *Nlrp1* deficiency (Fig. 3j). Interestingly, the biallelic loss of *Il-18* did not rescue DPP9 loss of function (Fig. 3j) suggesting that *Il-1* may be the main driver of the disease. Rescued mice exhibited a similar phenotype to *Dpp9*^*S729A/S729A*^ *Nlrp1*^*-/-*^ mice, appearing healthy yet runted. For example, *Dpp9*^*S729A/S729A*^ *Gsdmd*^*-/-*^ mice did not gain weight at the same rate as controls, and developed the same immune cell infiltrate in their lungs as *Dpp9*^*S729A/S729A*^ *Nlrp1*^*-/-*^ mice at 6 months of age, despite normal blood cell counts (Extended Data Fig. 4). The number of rescued *Dpp9*^*S729A/S729A*^ mice that survived to weaning age fell significantly below Mendelian ratios (Fig. 3j). However, these numbers might be confounded by potential cannibalisation of the runted pups. Overall it is clear that the deletion of a single allele of either *Asc, Gsdmd* or *Il-1r* is sufficient to protect *Dpp9*^*S729A/S729A*^ mice. These results raise the possibility that even partial inhibition of the NLRP1-inflammasome pathway, with anti-IL-1β antibodies for example, would provide some therapeutic benefit for DPP9 deficient patients. In further support of this, anti-IL-1β therapy was partially successful in a patient harbouring a germline NLRP1 p.P1214R mutation which abrogated binding and therefore inhibition by DPP9^21^.

In summary, we have identified two unrelated families with AR DPP9 deficiency, which is characterised by immune-associated defects, dermatological anomalies and neurological deficits. This Mendelian disease bears some degree of resemblance with auto-immune disorders caused by NLRP1 activating mutations. In particular the AIADK-causing NLRP1 p.P1214R mutation which entirely abrogates DPP9 binding and inhibition of NLRP1, is expected to yield comparable phenotypes^7^. Both diseases appear to manifest with recurrent episodes of fevers, auto-immune anemia and a general failure to thrive (Table 1). The milder skin manifestations seen in DPP9 patients may be explained by the presence of active DPP8 which we have shown can compensate for DPP9 deficiency in human cells^7^.

Using biochemical and cellular assays, we demonstrated that all patient-derived *DPP9* alleles had reduced catalytic activity and were defective in suppressing the NLRP1 inflammasome in vitro. These results suggest that the patients’ symptoms are at least in part due to the aberrant activation of the NLRP1 inflammasome. This notion was validated by in vivo experiments: the neonatal lethality of *Dpp9* mutant mice was rescued by the deletion of *Nlrp1* and downstream inflammasome signaling components *Asc, Gsdmd* and *Il-r1*. Thus IL-1β inhibition might be a viable therapeutic strategy for these patients. Recent studies have shown that human DPP9 has many cellular substrates and also represses an orthologous inflammasome sensor, CARD8, which is absent in rodents^22–26^. These findings could account for the broad spectrum of symptoms in patients with DPP9 mutations, some of which might not be modelled in mice. Overall, our results delineate a new human Mendelian disease of germline DPP9 deficiency, whose pathogenesis is largely driven by the aberrant activation of the NLRP1 inflammasome.

## Methods

### Human study participants

The study included 2 affected children from 2 distinct families. Both are affected with recessive germline *DPP9* mutations. The study protocol was approved by A*STAR IRB (2019-087) and genetic analyses were performed in accordance with bioethics rules of national laws of The Rockefeller University Hospital, New York, USA; Stanford University School of Medicine, Stanford, California, USA; and Necker-Enfants Malades Hospital, Paris, France. Informed consent was obtained for all patient and healthy control volunteers reported in the study.

### DNA extraction, quantification, and quality control

Genomic DNA of the affected individuals and other available family members was extracted from either whole blood using a variety of extraction protocols. DNA concentration and quality were assessed using NanoDrop (Thermo Scientific) and Qubit (Life technologies) fluorometers. A260/A280 ratios of 1.8 to 2.0 and A260/A230 ratios >1.5 were accepted. DNA fragmentation was assessed using agarose gel (0.8%) electrophoresis.

### Exome sequencing

Exome sequencing was employed independently for the detection of variants in Families 1 and 2.

Family 1 - Trio exome sequencing was performed using DNA extracted from peripheral blood samples at the Baylor Medical Genetics Laboratories according to published methods^27,28^. Briefly, capture was performed with VCRome 2.1 in-solution exome probes, as well as additional probes for over 3,600 Mendelian-disease-related genes. Library DNA was sequenced on an Illumina HiSeq for 100 bp paired-end reads. Data analysis was performed with Mercury 1.0. Genes possibly associated with the patient’s phenotype under a recessive mode of inheritance were reported in *DPP9, MS4A2, MXRA5* and *PAPLN*.

Family 2 - Genomic DNA (gDNA) was prepared from blood samples from patient II:1 by the standard phenol-chloroform extraction method. WES was performed with the SureSelect Human All Exon 71 Mb kit (Agilent Technologies). Paired-end sequencing was performed on a HiSeq 2500 machine (Illumina) generating 100-base reads. We aligned the sequences with the GRCh37 reference build of the human genome with the Burrows-Wheeler aligner^29^. Downstream processing and variant calling were performed with the Genome Analysis Toolkit^30^, SAMtools^31^. Substitution and InDel calls were made with the GATK Unified Genotyper. All variants were annotated with annotation software developed in-house^32–35^. Blacklisting variants common in private cohorts but not in public databases optimizes human exome analysis^35^. Human Gene Mutation Database for any given gene^36^. Based on the high consanguinity in this patient (1.66%), exome sequencing was analysed assuming autosomal recessive inheritance by keeping all homozygous non-synonymous coding variants and essential splice site variants with a MAF <10^−2^ in different ethnic subpopulations.

### Isolation of human fibroblasts and keratinocytes

Primary human fibroblasts and keratinocytes from Family 2 proband and two unaffected parental controls were isolated from fresh skin biopsies. Briefly, biopsies were incubated in dispase overnight at 4°C to enable peeling of the epidermis from the dermal compartment. Keratinocytes from the epidermis were then isolated by mechanical disruption and grown on a 3t3 feeder layer. Fibroblasts were isolated by finely chopping up the dermis and incubating it overnight in collagenase D. The collagenase was then removed the next day and fibroblasts were allowed to migrate out of the dermal fragments.

### Cell culture

All 293T lines and derivatives were cultured in DMEM supplemented with 10% FBS. Any control primary fibroblasts or keratinocytes used as WT controls were obtained from the Skin Research Institute of Singapore (SRIS) Skin Cell Bank with informed consent and prior IRB approval.

### SDS PAGE western blot and ELISA

Protein lysate was quantified using Bradford assay and 20 ug of protein loaded unless stated otherwise. Western blotting was carried out by incubating the primary antibodies overnight in 3% Milk in TBST. All ELISA experiments were carried out according to manufacturer’s instructions.

### Luminex multiplex microbead-based immunoassay

Conditioned supernatants were collected for Luminex analysis using the ProcartaPlex, Human Customized 65-plex Panel (Thermo Fisher Scientific,) to measure the following targets: APRIL; BAFF; BLC; CD30; CD40L; ENA-78; Eotaxin; Eotaxin-2; Eotaxin-3; FGF-2; Fractalkine; G-CSF; GM-CSF; Gro a; HGF; IFN-a; IFN-g; IL-10; IL-12p70; IL-13; IL-15; IL-16; IL-17 A; IL-18; IL-1 A; IL-1 b; IL-2; IL-20; IL-21; IL-22; IL-23; IL-27; IL-2 R; IL-3; IL-31; IL-4; IL-5; IL-6; IL-7; IL-8; IL-9; IP-10; I-TAC; LIF; MCP-1; MCP-2; MCP-3; M-CSF; MDC; MIF; MIG; MIP1a; MIP-1 b; MIP-3 A; MMP-1; NGF beta; SCF; SDF-1 A; TNF b; TNF-a; TNF-R2; TRAIL; TSLP; TWEAK; VEGF-A.

Harvested supernatants and standards were incubated with fluorescent-coded magnetic beads pre-coated with respective antibodies in a black 96-well clear-bottom plate overnight at 4°C. After incubation, plates were washed 5 times with wash buffer (PBS with 1% BSA (Capricorn Scientific) and 0.05% Tween-20 (Promega)). Sample-antibody-bead complexes were incubated with Biotinylated detection antibodies for 1 hour and washed 5 times with wash buffer. Subsequently, Streptavidin-PE was added and incubated for another 30 mins. Plates were washed 5 times again, before sample-antibody-bead complexes were re-suspended in sheath fluid for acquisition on the FLEXMAP® 3D (Luminex) using xPONENT® 4.0 (Luminex) software. Data analysis was done on Bio-Plex Manager^™^ 6.1.1 (Bio-Rad). Standard curves were generated with a 5-PL (5-parameter logistic) algorithm, reporting values for both mean fluorescence intensity (MFI) and concentration data. Hierarchical clustering and generation of heatmap was performed using pheatmap package on RStudio 1.3.1093.

### Mice

DPP9^S729A/S729A^ mice were provided by Prof Mark Gorrell (Centenary Institute). Nlrp1^-/- 37^, Gsdmd^-/- 38^, Ilr-1^-/- 39^, Asc^-/- 40^, Il-18^-/- 41^ were generated on, or backcrossed at least 10 generations onto the C57BL/6 background. Experiments use littermate controls where possible. All animal experiments complied with the regulatory standards of, and were approved by, the Walter and Eliza Hall Institute Animal Ethics Committee.

### Histology

Organs were collected in 10% neutral buffered formalin. Organ sections were prepared from paraffin blocks and stained with hematoxylin and eosin. Immunohistochemistry was performed using antibodies against F4/80 (in house), B220 (in house) or CD3 (Agilent #A045229).

### Micro-CT

For Micro-CT scans mice were euthanised and scanned on a Bruker Skyskan 1276 Micro-CT. Images were analysed using Amaris software.

### Hematology

Automated cell counts were performed on blood collected from the sub-mandibular vein into Microtainer tubes containing EDTA (Sarstedt), using an Advia2120i hematological analyser (Siemens, Munich, Germany).

### FACs analysis of newborn pup blood and bone marrow

Pups were euthanised 3-6 hours after birth via decapitation and blood collected from the neck into Microtainer tubes containing EDTA (Sarstedt). Limbs and sternum were collected, bones were crushed and passed through a 40 uM filter to obtain a single cell suspension. 1×10^6^ cells of the bone suspension and 15 uL of blood were plated per sample before red cell depletion using RCR Buffer (156 mM NH_4_Cl, 11.9 mM NaHCO_3,_ 0.097 mM EDTA). Cells were washed in PBS and incubated with rat anti-mouse CD16/CD23 (1:200 BioLegend #101302) at 4°C for 10 minutes to block Fc receptors. Cells were washed and stained for 1 hour at 4°C with fluorescently tagged antibodies: MHCII-FITC (1:200 in house), CD11c-PE (1:200 BioLegend #117307), CD4-PE-Cy7 (1:200 BioLegend #100528), CD3-PE-CF594 (1:200 BD Bioscience #562286), Ly6G-APC (1:200 in house), CD19-AF700 (1:200 BioLegend #15528), NK1.1-BV450 (1:400 BioLegend #108731), Zombie aqua viability Dye (1:500 Biolegend #77143), CD11b-BV605 (1:400 Biolegend 101257), CD8-BV650 (1:400 BioLegend #100742), Ly6C-BV711 (1:400 Biolegend #128037), CD45-BV786 (1:200 BioLegend #103149). Cells were washed and fixed with BD Phosflow Lyse/Fix buffer (BD Bioscience) for 20 min at RT and then stored at 4°C. Flow cytometry was performed on the BD LSRFortessa X-20 (BD Bioscience) and data acquired using BD FACSDiva (BD Bioscience) software. Data was analysed using FloJo (Tree Star). All cell populations shown were first gated to exclude small cellular debris and doublets, then gated on live cells (zombie negative) and CD45 positive, before gating on markers as shown in figure.

## Data Availability

The data that support the findings of this study are available from the corresponding
authors upon request.

## Acknowledgments

We are grateful to all members of the Masters, Zhong and Reversade laboratories for support. We thank Prof. Anthony Oro (Stanford University) for generous help in fibroblast derivation from family 1. We thank E. Kravets for study coordination and M.W. Allain for data collection (Stanford University). We thank S. Russo for outstanding animal husbandry. S.L.M. is supported by Australian National Health and Medical Research Council (NHMRC) Project Grants (2003159, 2003756) and Fellowships from the Victorian Endowment for Science Knowledge and Innovation, HHMI-Wellcome International Research Scholarship, and the Sylvia and Charles Viertel Foundation. The Laboratory of Human Genetics of Infectious Diseases is supported by the National Center for Research Resources and the National Center for Advancing Sciences (NCATS) of the National Institutes of Health (NIH) Clinical and Translational Science Award (CTSA) program (UL1TR001866), the French National Research Agency (ANR) under the “Investments for the Future” ANR program (ANR-10-IAHU-01), the Integrative Biology of Emerging Infectious Diseases Laboratory of Excellence (ANR-10-LABX-62-IBEID), and the “PNEUMOPID” project (Grant ANR 14-CE15-0009-01), the French Foundation for Medical Research (FRM) (EQU201903007798), the Howard Hughes Medical Institute, the Rockefeller University, the St. Giles Foundation, Institut National de la Santé et de la Recherche Médicale (INSERM), and the “Université de Paris”. K.L. is supported by a NMRC Open Fund - Young Individual Research Grant (OF-YIRG19MAY-0039). S.D. is supported by the National Health and Medical Research Council Early Career Fellowship (GNT1143412). C.-H.Y. is supported by the WEHI Centenary Fellowship and Ormond College’s Thwaites Gutch Fellowship in Physiology. F.L.Z. is an Nanyang Assistant Professor and a recipient of National Research Foundation (NRF, Singapore) fellowship. B.R. is an investigator of the National Research Foundation (NRF, Singapore) and Branco Weiss Foundation (Switzerland) and an EMBO Young Investigator, and is supported by a Use-Inspired Basic Research (UIBR) funding scheme and an inaugural Agency for Science, Technology and Research (A*STAR) Investigatorship (Singapore).

## Author contributions

F.L.Z, S.L.M, and B.R designed the study. J.B., D.B., H.K., N, N and N.H. made clinical diagnoses and collected clinical data and samples. C.H, P.L, S.D, C.-H.Y and S.L.M performed and supervised the mouse experiments. K.R., R.M.T and K.L. performed biochemical and cell-based experiments and analyzed the data. C.H, K.R, K. L, F.L.Z, S.L.M and B.R wrote the manuscript with input from all authors.

---

**Extended Data Fig. 1:**
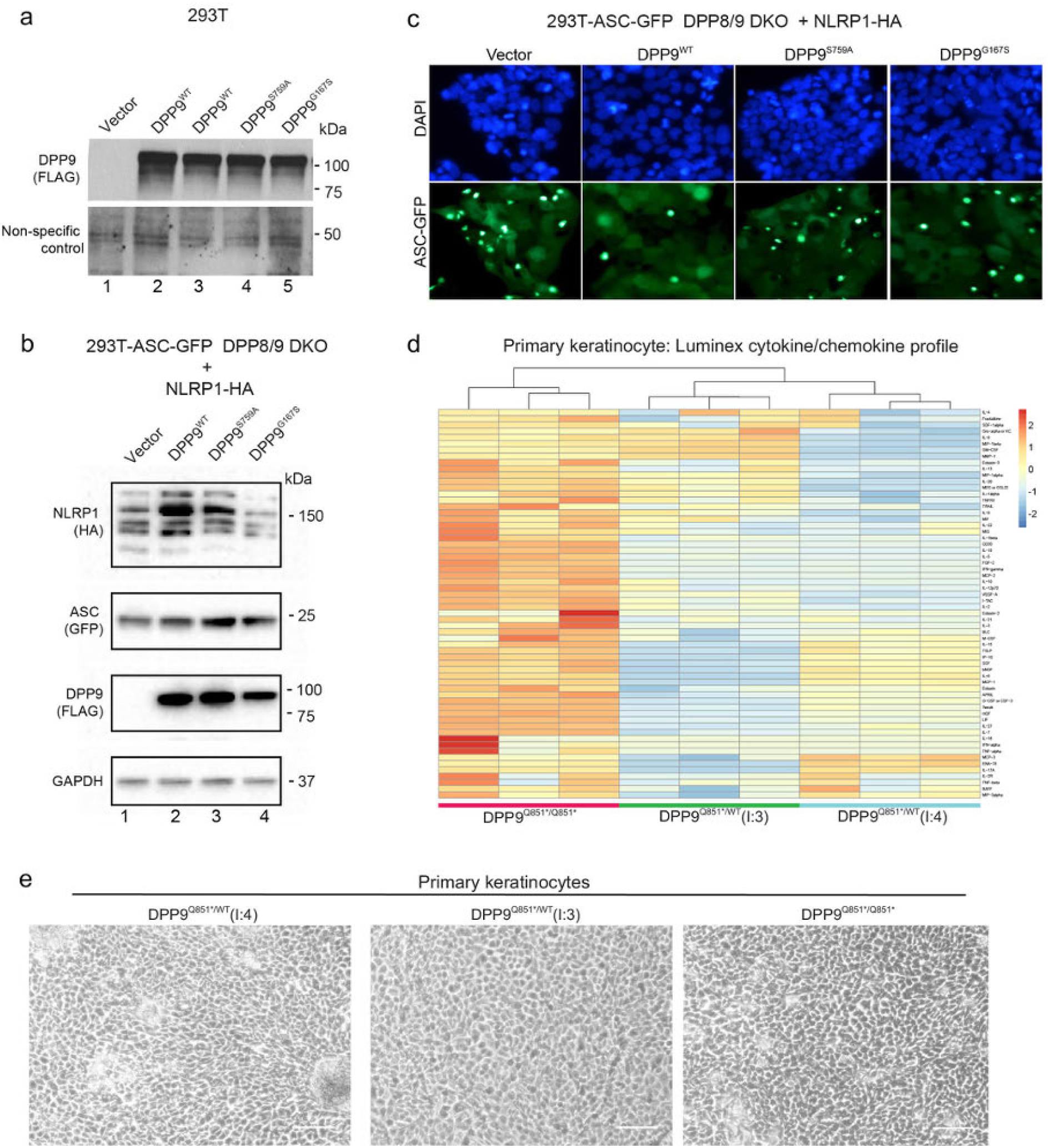
Mutant DPP9 alleles fail to inhibit NLRP1, eliciting release of proinflammatory cytokines. **a**, DPP8/9 enzyme assay shows a reduction in DPP8/9-derived enzymatic activity in DPP9 S759A and G167S mutants. Western blot analysis of DPP9 protein levels or a non-specific loading control for data obtained in Fig. 2b. **b**, DPP9 S759A and G167S mutants are unable to repress NLRP1 inflammasome. Western blot analysis for NLRP1-HA, ASC-GFP and FLAG-DPP9, to confirm their expression in DPP8/9 DKO cells used in Fig. 2c. **c**, DPP9 S759A and G167S mutants are unable to repress NLRP1 inflammasome. 293T-ASC-GFP DPP8/9 DKO cells were transfected with a vector control or WT NLRP1 together with wild-type DPP9, DPP9 S759A, or DPP9 G167S. Figure shows representative images of percentage of cells with ASC-GFP specks for each transfected sample. **d**, Heatmap depicting Luminex cytokine/chemokine analysis and hierarchical clustering of primary keratinocyte cultures derived from proband and heterozygous parents of family 2. **e**, Primary keratinocytes isolated from the family 2 proband are morphologically indistinguishable from the unaffected parents. Phase contrast images of primary keratinocytes from the two unaffected parents from family 2 ((I:4) and (I:3) Q851X/+) and affected individual from family 2 (proband (II:4) Q851X/Q851X).

**Extended Data Fig. 2:**
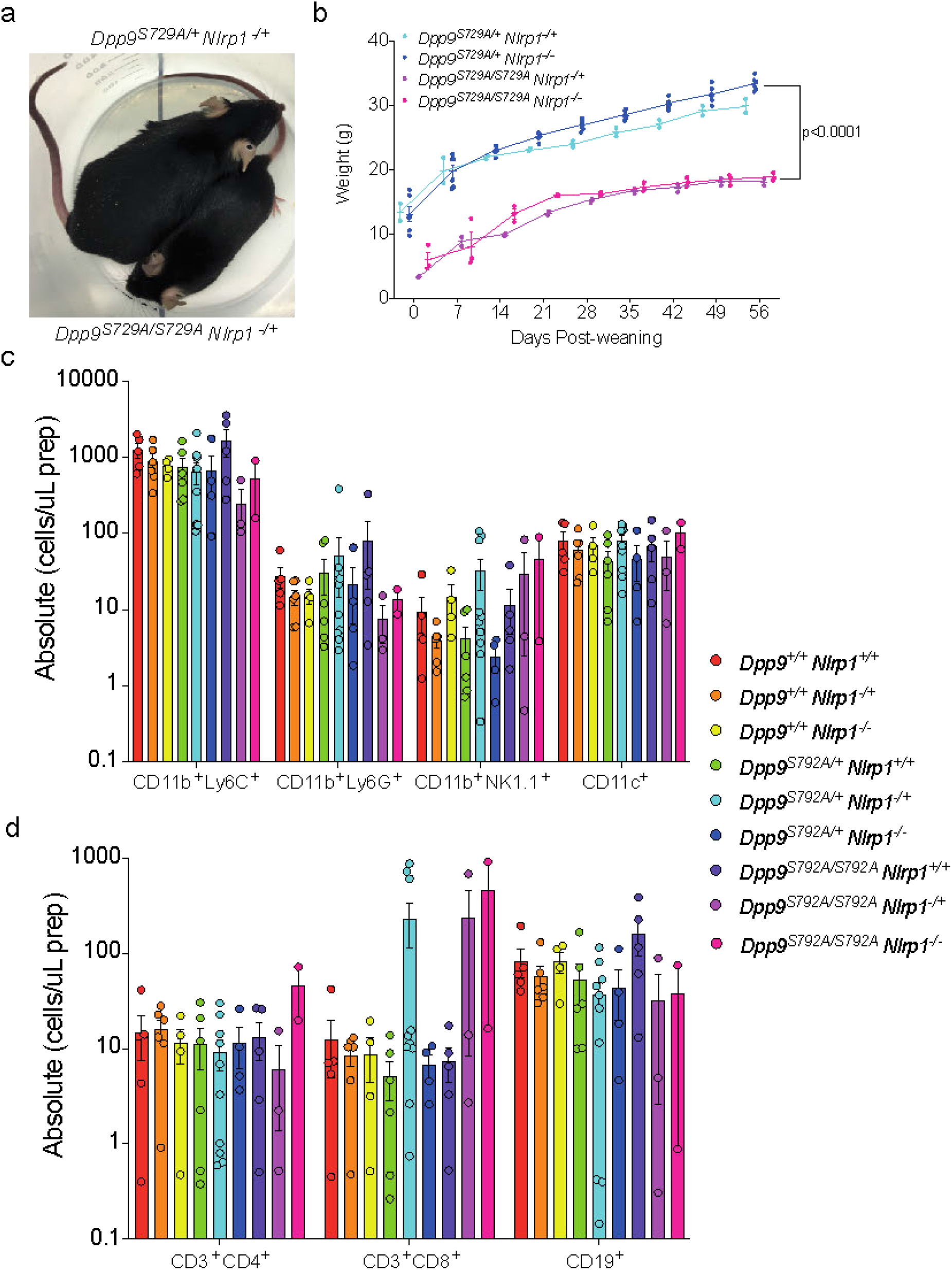
Removing one copy of NLRP1 is sufficient to rescue neonatal lethality of *Dpp9*^*S729A/S729A*^ mice. **a**, A viable, male *Dpp9*^*S729A/S729A*^ *Nlrp1*^*-/+*^ mouse is runted compared to a *Dpp9*^*S729A/+*^ *Nlrp1*^*-/+*^ littermate. **b**, *Dpp9*^*S729A/S729A*^ *Nlrp1*^*+/-*^ mice exhibit stunted weight gain compared to *Dpp9*^*S729A/+*^ *Nlrp1*^*+/-*^ controls. Male *Dpp9*^*S729A/S729A*^ *Nlrp1*^*-/+*^ and *Dpp9*^*S729A/+*^ *Nlrp1*^*-/+*^ mice were weighed weekly for 8 weeks post weaning (n=2-5 per genotype). **c, d**, FACs analysis of immune cells of bone marrow collected from 3-6 hour-old pups. Error bars represent mean ±SEM. Statistical significance was determined by one-way ANOVA.

**Extended Data Fig. 3:**
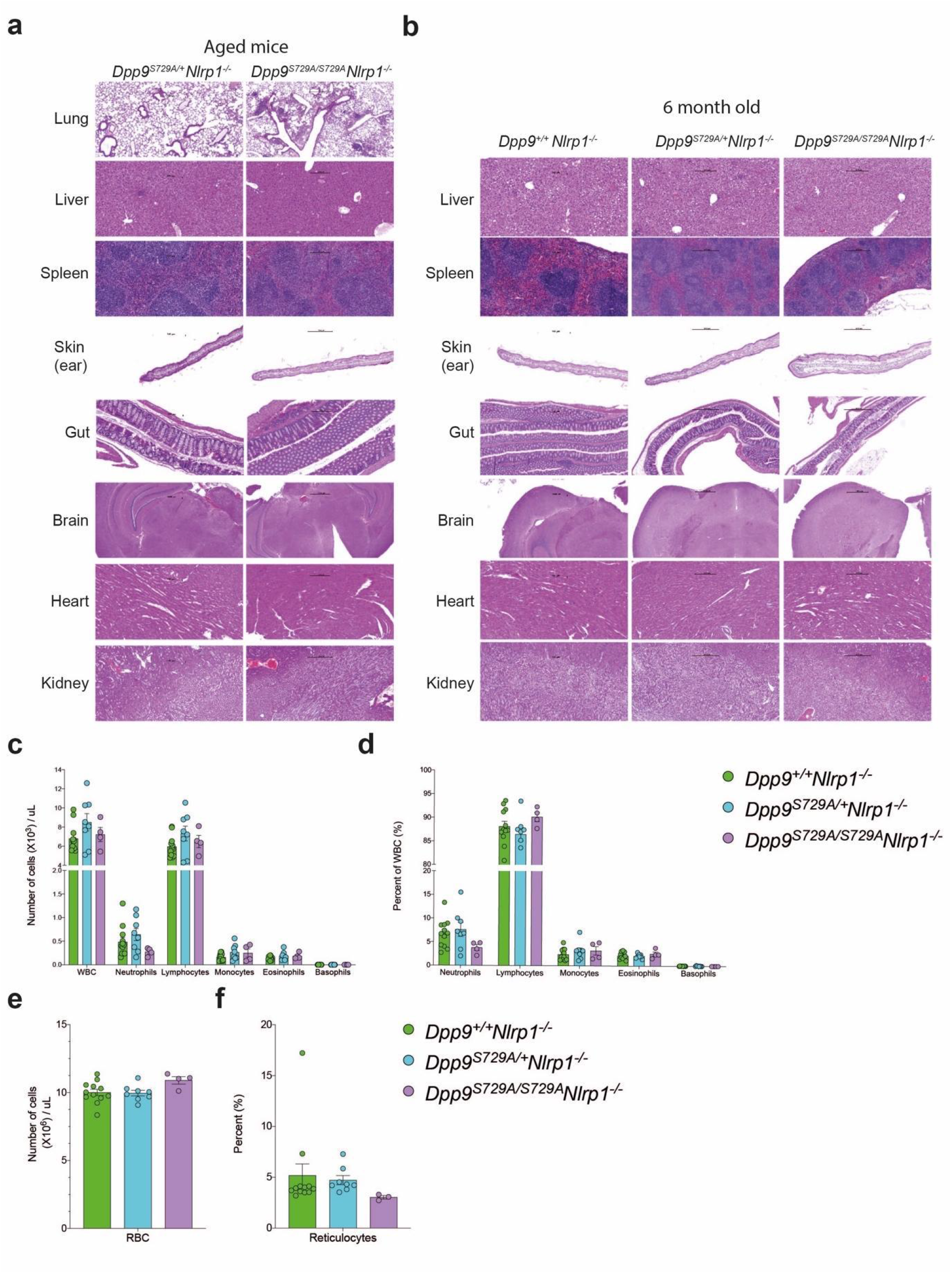
*Dpp9*^*S729A/S729A*^ *Nlrp1*^*-/-*^ mice display accumulation of immune cell infiltrate in the lung. **a**, *Dpp9*^*S729A/S729A*^ *Nlrp1*^*-/-*^ display immune cell infiltrate in lungs. Hematoxylin and eosin stained section of lung, liver, spleen, sternum, skin, gut, brain, heart and kidney tissue from aged and **b**, 6-month-old *Dpp9*^*S729A/S729A*^ *Nlrp1*^*-/-*^ and *Dpp9*^*S729A/+*^ *Nlrp1*^*-/-*^ mice. Black arrows indicate immune cell infiltrate. N=3-7 per genotype. **c-f**, Peripheral blood cell numbers remain unchanged in adult (4-months-old) *Dpp9*^*S729A/S729A*^ *Nlrp1*^*-/-*^ mice compared to controls. Absolute **(c)** and percentage **(d)** of neutrophils, lymphocytes, monocytes, eosinophils and basophils. **e**, Absolute number of red blood cells. **f**, Percentage of reticulocytes. Error bars represent mean ±SEM.

**Extended Data Fig. 4:**
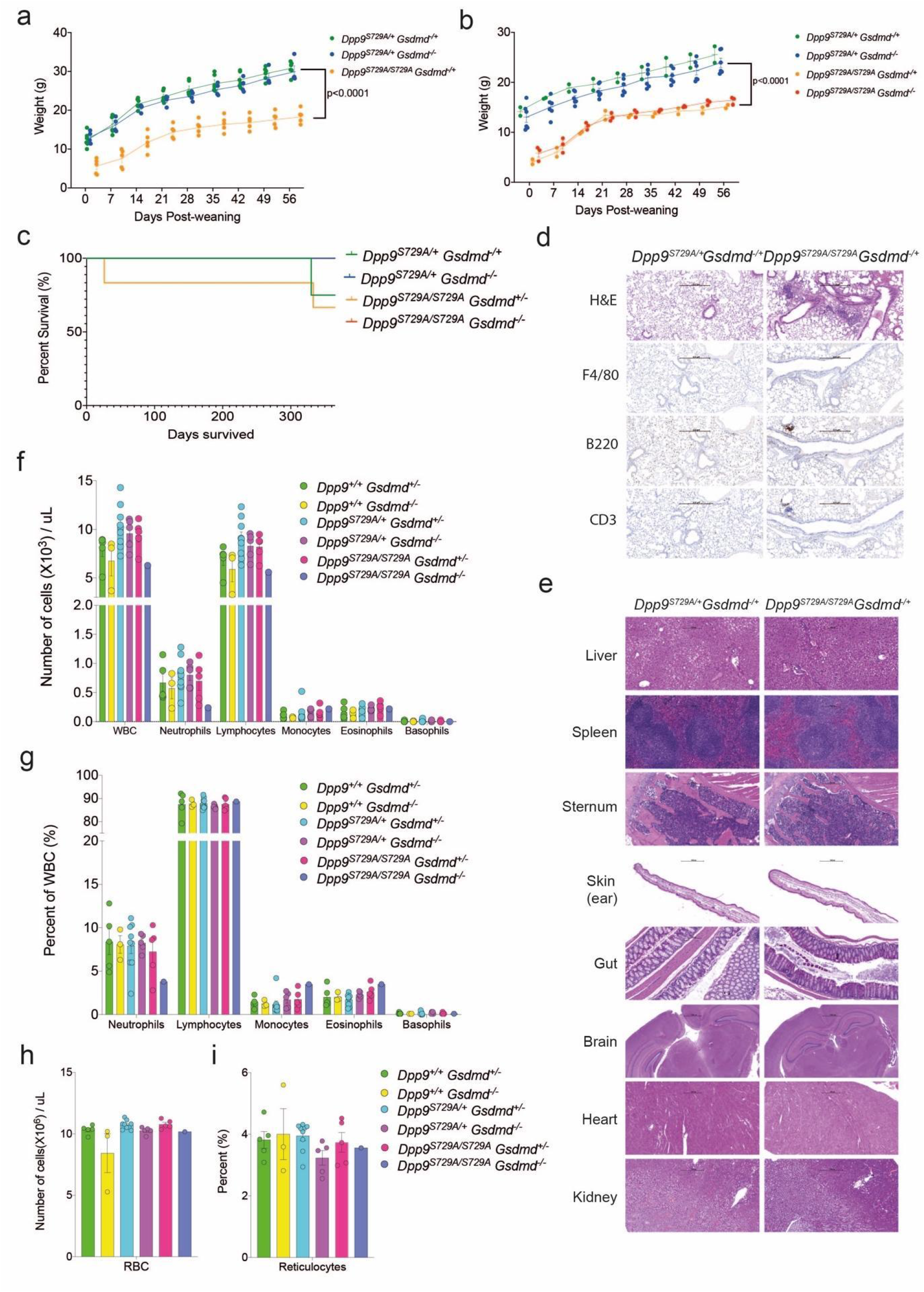
*Dpp9*^*S729A/S729A*^ *Gsdmd*^*-/-*^ mice phenocopy *Dpp9*^*S729A/S729A*^ *Nlrp1*^*-/-*^ mice. **a**, Male and **b**, female *Dpp9*^*S729A/S729A*^ deficient in GSDMD are runted compared to *Dpp9*^*S729A/+*^ mice deficient in GSDMD. Mice were weighed weekly for 8 weeks post weaning to quantify the size difference. **c**, Post-weaning Kaplan Meier survival curve of *Dpp9*^*S729A/S729A*^ or *Dpp9*^*S729A/+*^ mice deficient in GSDMD showing no difference in survival up to 300 days. **d**, *Dpp9*^*S729A/S729A*^ *Gsdmd*^*-/+*^ mice display immune infiltrate in the lungs. Immunohistochemistry of F4/80, B220 or CD3 antibodies on sections of mouse lung and **e**, hematoxylin and eosin stained and on section of lung, liver, spleen, sternum, skin, gut, brain, heart and kidney tissue from 6-month-old *Dpp9*^*S729A/S729A*^ *Gsdmd*^*-/+*^ and *Dpp9*^*S729A/+*^ *Gsdmd*^*-/+*^ mice (n=2-4 per genotype). **f-i**, Peripheral blood cell numbers remain unchanged in adult (4-months-old) *Dpp9*^*S729A/S729A*^ *Gsdmd*^*-/-*^ mice compared to controls. Absolute **(f)** and percentage **(g)** of neutrophils, lymphocytes, monocytes, eosinophils and basophils. **h**, Absolute number of red blood cells. **i**, Percentage of reticulocytes. Error bars represent mean ±SEM. Statistical significance was determined by one-way ANOVA.

